# Performance of a diagnostic model for the presence of unruptured intracranial aneurysms in the general population

**DOI:** 10.1101/2023.04.19.23288842

**Authors:** Vita M. Klieverik, Bob Roozenbeek, Tim Y. Cras, Meike W. Vernooij, Mirjam I. Geerlings, Daniel Bos, Ynte M. Ruigrok

## Abstract

**Background:** The prevalence of unruptured intracranial aneurysms (UIAs) in the general population is 3%. Aneurysmal subarachnoid hemorrhage can be prevented by screening for UIAs followed by preventive treatment of identified UIAs. We developed a diagnostic model for presence of UIAs in the general population to help identify persons at high risk of having UIAs.

**Methods:** Between 2005-2015, middle aged and elderly participants from the population-based Rotterdam Study underwent brain magnetic resonance imaging at 1.5 Tesla, on which presence of incidental UIAs was evaluated. A multivariable logistic regression model with diagnostic markers sex, age, hypertension, smoking, hypercholesterolemia, diabetes, and alcohol, and their interactions, was developed. We corrected for overfitting using bootstrapping. Model performance was assessed with discrimination, calibration, sensitivity, specificity, positive predictive value (PPV), and negative predictive value (NPV).

**Results:** 5835 persons were included (55.0% women, mean age 64.9±10.9 years) with a 2.2% UIA prevalence. Sex, age, hypertension, smoking, diabetes, and interactions of sex with age, hypertension, and smoking were independent diagnostic markers. The resulting model had a c-statistic of 0.65 (95% confidence interval [CI] 0.60–0.68) and 56% sensitivity, 52% specificity, 98% PPV, and 3% NPV for UIA presence at a cut-off value of 4% prevalence. Because of interactions with sex, additional models for men and women separately were developed. The model for men had a c-statistic of 0.70 (95%CI 0.62–0.78) with age, hypertension, and smoking as diagnostic markers and comparable additional performance values as for the full model. The model for women had a c-statistic of 0.58 (95%CI 0.52–0.63) with smoking as the only diagnostic marker.

**Conclusion:** Our diagnostic model had insufficient performance to help identify persons at high risk of having UIAs in the general population. Rather, it provides insight in risk factors contributing to UIA risk and shows that these may be in part sex-specific.

## Introduction

Unruptured intracranial aneurysms (UIAs) are fairly common with a 3% prevalence in the general population.^1^ Rupture of an aneurysm results in aneurysmal subarachnoid hemorrhage (aSAH), a severe subtype of stroke with an incidence of 6 per 100 000 person years, resulting in a lifetime risk of around 0.5%.^2,3^ Established major risk factors for both UIAs and aSAH are female sex, hypertension, smoking and a positive family history for aSAH.^1,4^ Around one-third of patients with aSAH die, and of those who survive, one half remains dependent on continuous care of others.^5^ Although the other half of patients regains independence, 95% of them suffer from severe cognitive impairments, which greatly affect their functionality and quality of life.^6^ Approximately 12% of patients with aSAH die before receiving medical attention and for those patients admitted to hospital, the early effects of aSAH are the principal cause of death.^7,8^ Thus, prevention of aSAH has high potential to prevent poor outcome from aSAH.^9^ As UIAs are hardly ever symptomatic before they rupture, screening is the only way to detect UIAs before rupture, and to install preventive treatment.

The prevalence of UIAs is approximately three times higher in persons with a family history of aSAH compared to persons without such a family history.^1^ Moreover, their risk of developing aSAH doubles in case of one affected first-degree relative and increases fifty times in case of two or more affected relatives which confers to a lifetime risk of around 25%.^10^ Repeated radiological screening for early detection of UIAs in persons with two or more affected first-degree relatives has shown to be cost-effective, with newly identified UIAs at first screening in 11% and in 8% at follow-up screening.^11,12^

As a positive family history for aSAH accounts for around 10% of aSAH cases,^4,13^ screening and preventive treatment of patients with familial preponderance of aSAH alone will only cause a modest reduction of aSAH incidence at a population level. Therefore, additional high-risk individuals within the general population in whom screening might also be effective, should be identified. As a first step in this identification, it is important to know who in the general population are at high risk of UIAs.

Therefore, the aim of this study is to develop a diagnostic model for the detection of UIAs in the general population to help identify persons at high risk of having an UIA.

## Methods

This study was performed in accordance with the Transparent Reporting of a Multivariable Prediction Model for Individual Prognosis or Diagnosis (TRIPOD) statement, a set of recommendations for the reporting of studies developing and validating a prediction model.^14,15^

### Study design and study population

For this cross-sectional study, we used data from the Rotterdam Study, a general population-based prospective cohort study containing over 14 000 participants aged 45 years or older and recruited from 1990 onwards in Ommoord, a neighborhood in Rotterdam, the Netherlands.^16^ These participants attended a dedicated study center for baseline and follow-up assessments, where they were invited to fill out questionnaires, had physical measurements taken, provided biological samples, and underwent medical imaging.^16^ For our study we used data from participants of a subset of the Rotterdam Study who had undergone brain magnetic resonance imaging (MRI) scanning between 2005 and 2015.^17^ The Rotterdam Study has obtained approval from the Medical Research Ethics Committee (MREC) of the Erasmus MC University Medical Center Rotterdam (registration number MEC 02.1015) and from the review board of the Dutch Ministry of Health, Welfare and Sports (Population Screening Act WBO, license number 10712172-159521-PG) and this approval is renewed every five years.^16^ The Rotterdam Study is registered into the Netherlands National Trial Register and the World Health Organization (WHO) International Clinical Trials Registry Platform (shared catalogue number NTR6831).^16^ All participants provided their written informed consent prior to participating in the Rotterdam Study.

### Assessment of UIAs

Our primary outcome measure was prevalent UIA on proton-density T2-weighted brain MRI scans for which a study-dedicated 1.5 Tesla MRI scanner with an 8-channel head coil (General Electric Healthcare, Milwaukee, USA) was used.^17^ The brain MRI scan protocol has been described elsewhere.^17^ Per study protocol, each brain MRI scan was assessed by trained research physicians for the presence of incidental findings of potential clinical relevance, including UIAs.^18,19^ All UIAs detected by the research physicians were subsequently reviewed by experienced neuroradiologists including assessment of location and size of the UIAs.^18^ Discrepancies in the assessment of the research physicians and neuroradiologists were discussed and solved in a consensus meeting.^18^ In accordance with the study protocol on management of incidental findings, all participants with an UIA larger than seven millimeters in diameter in the anterior circulation or an UIA in the posterior circulation were referred for additional clinical assessment.^18,19^ Only participants with saccular UIAs were included and patients with fusiform UIAs were thus excluded, as fusiform UIAs have a distinctly different etiology and a much lower risk of aSAH.^4^

### Assessment of potential diagnostic markers

Prior examination of the literature guided the selection of candidate diagnostic markers, which were limited to those that are routinely available or can be easily ascertained by general practitioners during a standard consultation. These included sex, age, hypertension, smoking status, hypercholesterolemia, diabetes mellitus (DM), and alcohol consumption.^18,20,21^ The interaction between smoking status and alcohol consumption was included since these candidate diagnostic markers were expected to influence their mutual associations with prevalent UIA. In addition, we included interactions between sex and diagnostic markers to study sex as a potential effect modifier for the following reasons: 1. the female preponderance of the disease with two-thirds of patients being women^1,4^ and 2. suggested differential effects according to sex of risk factors.^22^ All candidate diagnostic markers were assessed at the time of brain MRI scanning.^18^ Hypertension was defined as systolic blood pressure ≥ 140 mmHg or diastolic blood pressure ≥ 90 mmHg and/or the use of antihypertensive medication. Blood pressure was assessed two times using a random zero sphygmomanometer, of which the mean value was taken.^18^ Smoking status was grouped into (1) never smokers, (2) former smokers, and (3) current smokers. Hypercholesterolemia was defined as a total cholesterol level of ≥ 6.2 mmol/l and/or the use of cholesterol lowering medication.^18^ DM was defined as a fasting blood glucose level of > 7.0 mmol/l and/or a non-fasting blood glucose level of > 11.1 mmol/l and/or use of antidiabetic medication.^18^ Alcohol consumption was measured in grams per day, assuming one glass contains ten grams of alcohol.^18^ We categorized alcohol consumption into (1) no alcohol consumption, (2) alcohol consumption < 150 grams per week, and (3) alcohol consumption ≥ 150 grams per week, which was considered excessive alcohol consumption.^21^ Data on smoking status and alcohol consumption were acquired through home interviews.^17^

### Statistical analysis

Normally distributed continuous variables were presented as means ± standard deviations (SD), while skewed distributed continuous variables were expressed as medians with corresponding interquartile ranges (IQR). Categorical variables were shown as numbers with corresponding percentages. Data were missing for hypertension (0.5%), smoking status (0.6%), total cholesterol level (1.7%), cholesterol lowering medication (0.6%), DM (1.2%), and alcohol consumption (5.7%). Missing data were imputed using the expectation maximization algorithm based on the variables hypertension, smoking status, hypercholesterolemia, alcohol consumption, and UIA prevalence.^18,23^

To study the association between candidate diagnostic markers and prevalent UIAs, we developed a multivariable logistic regression model. We used restricted cubic splines to evaluate whether the continuous candidate diagnostic marker age could be analyzed as a linear variable or required transformation. To assess whether candidate diagnostic markers contributed to the model, we performed backward selection based on Akaike Information Criterion (AIC).^24^ Models derived from multivariable regression can be too optimistic and overestimate effect estimates when applied to a different population.^25^ To correct for this, we internally validated the model by applying a shrinkage factor to the regression coefficients, determined by bootstrapping procedures.^25^ The estimated effect sizes of the independent diagnostic markers derived from the model was expressed as odds ratio’s (OR) with corresponding 95% confidence intervals (CI). As the model assessing the whole study population included several interactions between sex and other candidate diagnostic markers as independent markers, we decided to additionally develop diagnostic models for men and women separately.

We assessed the performance of the models by estimating their discrimination, calibration, sensitivity, specificity, positive predictive value (PPV), and negative predictive value (NPV).^25,26^ Discrimination refers to the models’ ability to correctly distinguish participants with an UIA from those without an UIA and we evaluated this ability using the concordance statistic (c-statistic).^25^ Calibration is an indicator for the measure of agreement between predicted and observed probability of prevalent UIAs and we assessed this visually with a calibration plot.^25^ We aimed to use a cut-off value of 4% for UIA prevalence for estimating sensitivity, specificity, PPV, and NPV in the model, meaning that persons who have a predicted risk of having an UIA of 4% or higher were classified as having a positive result. We based this cut-off value on the finding that screening for UIAs in persons with one first-degree relative who had an aSAH is likely to be cost-effective in case of an UIA prevalence of 4%.^29^ If in a model no individuals had a predicted UIA prevalence of 4% we used a cut-off value of 1% instead, to ensure there were enough UIAs to test the model’s performance with. All statistical analyses were performed using R statistical software, version 4.0.2 (R Foundation for Statistical Computing, Vienna, Austria). To calculate an individual person’s absolute risk of having an UIA, we provided the original regression equation of the diagnostic models.

## Results

### Baseline characteristics

After excluding participants with fusiform UIAs (n=6), 5835 participants were included in the current study (Table 1). Of these participants, 3211 (55.0%) were women and the mean age was 64.9 ± 10.9 years. A total of 130 (2.2%) participants had a saccular UIA, of whom 89 (68.5%) were women.

**Table 1.** Baseline characteristics of the study population

|  | All participants | Men | Women |
| --- | --- | --- | --- |
| Characteristic | (n = 5835) | (n = 2624) | (n = 3211) |
| Prevalent UIA | 130 (2.2) | 41 (1.6) | 89 (2.8) |
| Age (years) |  |  |  |
| < 50 | 428 (7.3) | 191 (7.3) | 238 (7.4) |
| ≥ 50 | 5407 (92.7) | 2433 (92.7) | 2973 (92.6) |
| Mean ± SD | 64.9 ± 10.9 | 64.7 ± 10.7 | 65.1 ± 11.0 |
| Hypertension | 3715 (63.7) | 1740 (66.3) | 1973 (61.4) |
| Smoking status |  |  |  |
| Never smoking | 1883 (32.3) | 635 (24.2) | 1246 (38.8) |
| Former smoking | 2956 (50.7) | 1533 (58.4) | 1424 (44.3) |
| Current smoking | 996 (17.1) | 456 (17.4) | 541 (16.8) |
| Hypercholesterolemia | 2406 (41.2) | 1045 (39.8) | 1362 (42.4) |
| DM | 765 (13.1) | 412 (15.7) | 353 (11.0) |
| Alcohol consumption |  |  |  |
| Never | 474 (8.1) | 154 (5.9) | 320 (10.0) |
| < 150 grams per week | 4840 (82.9) | 2083 (79.4) | 2757 (85.9) |
| ≥ 150 grams per week | 521 (8.9) | 387 (14.7) | 134 (4.2) |

Data are n (%), unless otherwise indicated. UIA = unruptured intracranial aneurysm, SD = standard deviation, DM = diabetes mellitus.

### Diagnostic models and performance

Table 2 shows the results from the logistic regression analysis of diagnostic markers of prevalent UIAs in the full study population. Restricted cubic splines showed that age could be analyzed as a continuous variable. Sex, age, hypertension, smoking status, DM, and the interactions of sex with age, hypertension, and smoking status were independent diagnostic markers of prevalent UIAs. Following internal validation using bootstrapping procedures, the c-statistic of this model was 0.65 (95% CI 0.60 – 0.68). The model had 56% sensitivity, 52% specificity, 98% PPV, and 3% NPV for UIA detection at a cut-off value of 4%. The calibration plot of observed and predicted probabilities showed that this model slightly overestimated UIA risk (Figure 1A).

**Table 2.** Univariable and multivariable logistic regression analysis of diagnostic markers of prevalent unruptured intracranial aneurysms (UIAs) in the full study population

|  | Univariable | Multivariable* |
| --- | --- | --- |
| Diagnostic marker | OR (95% CI) | OR (95% CI) |
| Female sex | 1.80 (1.24 – 2.61) | 14.43 (0.97 – 214.88) |
| Age per year | 1.01 (0.99 – 1.02) | 1.02 (0.99 – 1.06) |
| Hypertension | 1.45 (0.98 – 2.12) | 2.38 (0.95 – 5.99) |
| Smoking status |  |  |
| Never smoking | Reference |  |
| Former smoking | 1.54 (0.96 – 2.46) | 1.43 (0.46 – 4.39) |
| Current smoking | 3.52 (2.14 – 5.77) | 4.67 (1.51 – 14.39) |
| DM | 0.67 (0.37 – 1.22) | 0.70 (0.38 – 1.29) |
| Interactions |  |  |
| Female sex*Age per year | 0.97 (0.94 – 1.00) | 0.98 (0.94 – 1.02) |
| Female sex*Hypertension | 0.39 (0.15 – 1.04) | 0.52 (0.18 – 1.47) |
| Female sex*Former smoking | 0.87 (0.26 – 2.92) | 1.07 (0.31 – 3.72) |
| Female sex*Current smoking | 0.47 (0.14 – 1.63) | 0.52 (0.15 – 1.87) |

\* The initial regression coefficients were corrected for overfitting with a shrinkage factor of 0.68. OR
= odds ratio, CI = confidence interval, DM = diabetes mellitus.

**Figure 1.**
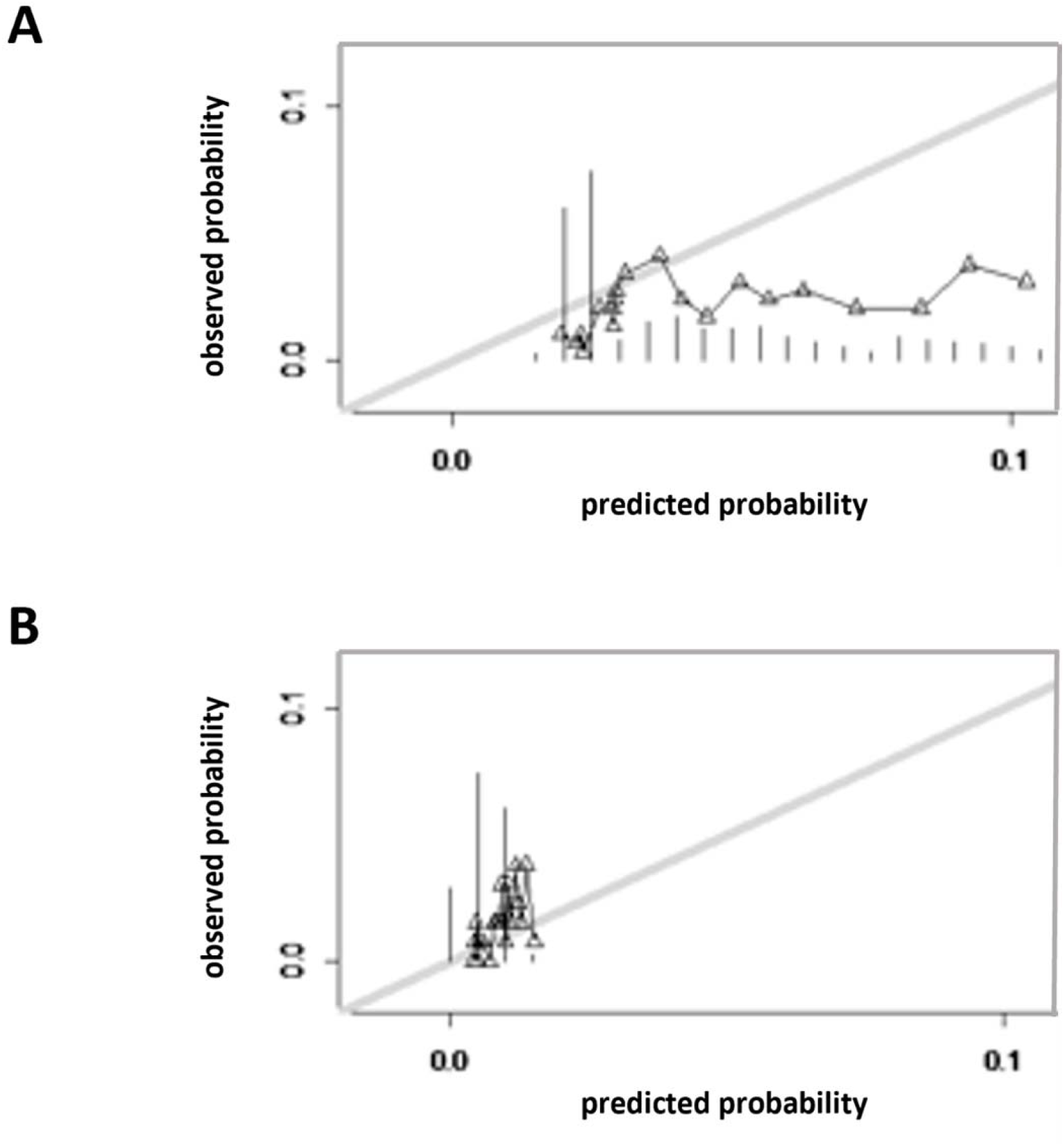
Calibration plot of the diagnostic model for the full study population (panel A) and for men only (panel B)

Table 3 shows the results from the model for men and women only. Age, hypertension, and smoking status were independent diagnostic markers of prevalent UIAs in men. The c-statistic was 0.70 (95% CI 0.62 – 0.78). In this model no individuals had a predicted UIA prevalence of 4% so we used a cut-off value of 1% for UIA prevalence instead for further assessing the performance of the model. The model had 62% sensitivity, 55% specificity, 99% PPV, and 2% NPV for UIA detection at a cut-off value of 1%. The calibration plot demonstrated a fair agreement between the observed and predicted probability of prevalent UIAs (Figure 1B).

**Table 3.**
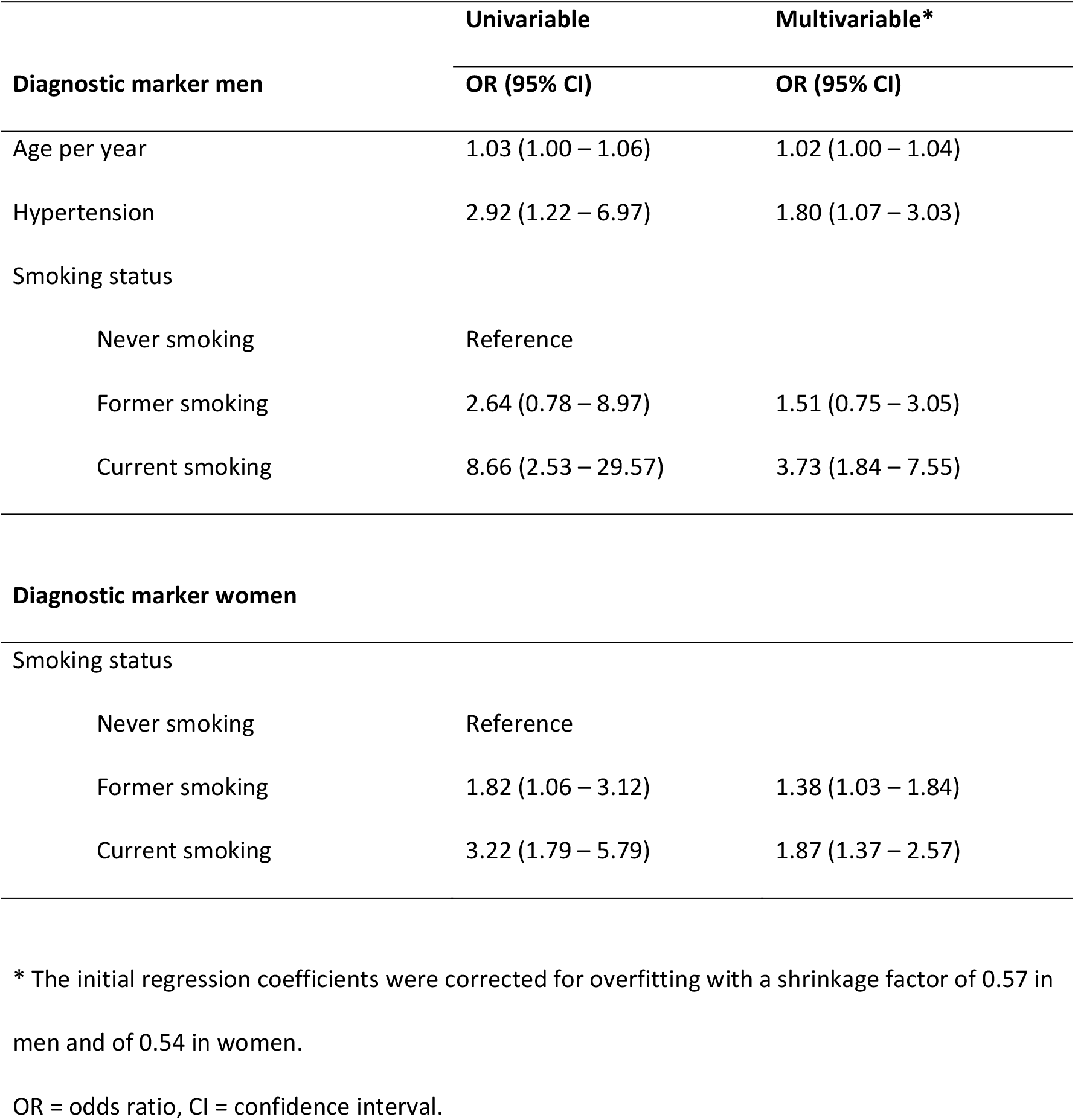
Univariable and multivariable logistic regression analysis of diagnostic markers of prevalent unruptured intracranial aneurysms (UIAs) in men and women separately

Smoking status was the only independent diagnostic marker of prevalent UIAs in women. The c-statistic of this model was 0.58 (95% CI 0.52 – 0.63). As there was only one diagnostic marker in this model we were unable to calculate additional performance measures for this model.

### Individual absolute risks of having an UIA

The original regression equation of the diagnostic models for the whole population and for men only are provided in Figure 2 and Figure 3, respectively. Based on the diagnostic model for the whole population, the risk of having an UIA ranged from 1.61% to 11.01%, while in the model for men only, this risk ranged from 0.39% to 1.86%.

**Figure 2.**
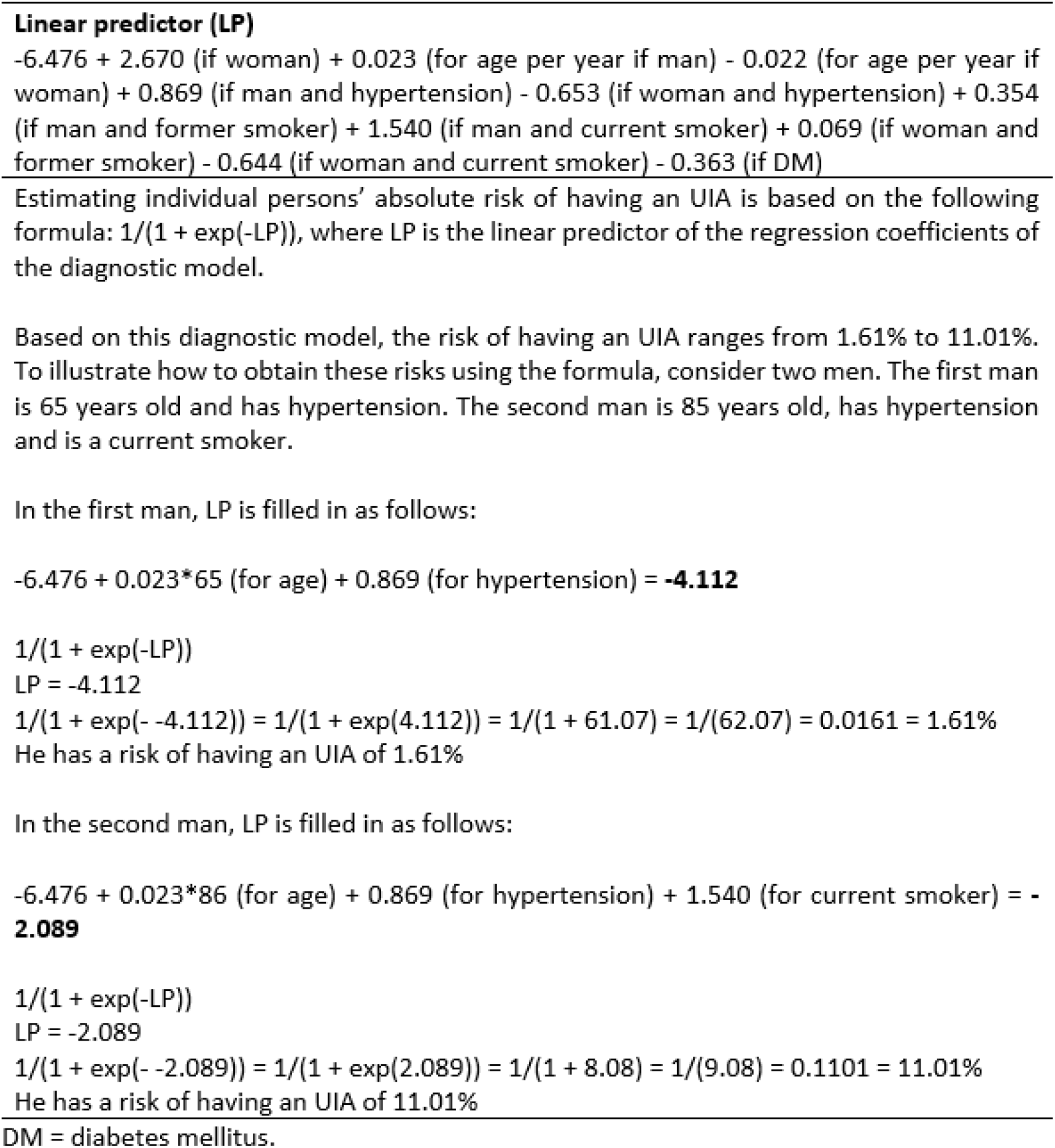
Original regression equation of the diagnostic model for the full study population to calculate the absolute risk of having an unruptured intracranial aneurysm (UIA)

**Figure 3.**
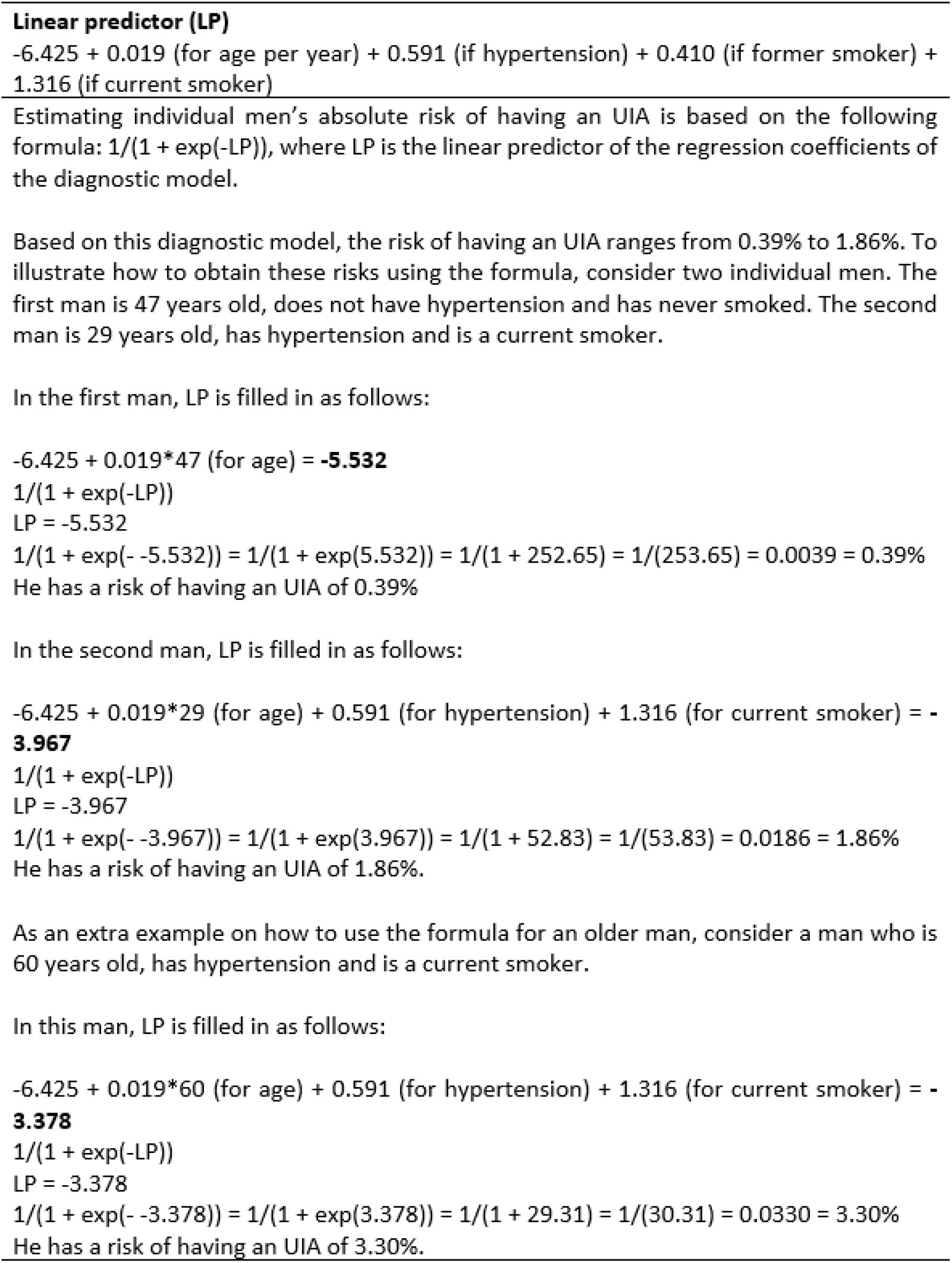
Original regression equation of the diagnostic model for men to calculate the absolute risk of having an unruptured intracranial aneurysm (UIA) for individual men

## Discussion

We developed three separate diagnostic models for estimating individual persons’ absolute risk of having an UIA in the general population (for the whole population, and for men and women separately) based on a set of person characteristics that are routinely available or easily ascertainable in a primary health care setting. We found that the model for men showed the best performance with a c-statistic of 0.70 (95% CI 0.62 – 0.78) with a fair agreement between the observed and predicted probability of prevalent UIAs. However, its additional performance values were suboptimal. Age, hypertension, and smoking status were confirmed as independent diagnostic markers of prevalent UIAs in men and the risk of having an UIA in men ranged from 0.39% to 1.86%. The model for the whole population had a somewhat lower performance with a c-statistic of 0.65 (95% CI 0.60 – 0.68) with the additional performance values being comparable to the values of the model for men. Next to sex, age, hypertension, smoking status and DM being independent diagnostic markers of prevalent UIAs in this model, three interactions of sex with other diagnostic markers (age, hypertension, and smoking status) were also identified as independent diagnostic markers. Last, the model for women performed the worst with a c-statistic of 0.58 (95% CI 0.52 – 0.63), and with smoking status identified as the only independent diagnostic marker of prevalent UIAs in women.

Since UIA rupture has devastating effects, false negative results for UIA detection should be as low as possible. Also, as in our study population the UIAs were diagnosed on proton-density T2-weighted brain MRI scans, instead of on magnetic resonance angiography (MRA), an additional MRA (or CT angiography) needs to be performed to confirm UIA presence. Therefore, specificity should preferably be also high to reduce costs and burden for patients. Consequently, a diagnostic model for UIA detection in the general population requires both high sensitivity and specificity.^26^ Although the model for men showed the best performance out of all three models, its sensitivity of 62% and specificity of 55% for UIA presence were still limited. The limited performance of our model may be explained by the relatively low prevalence of UIAs which contrasts with the relatively prevalent diagnostic markers for this disease, being sex, age, hypertension, smoking status, hypercholesterolemia, DM, and alcohol consumption.^18,20,21^ Moreover, in this model the upper limit of the risk of having an UIA was only 1.86% while the overall prevalence of UIAs in the general population is around 3%.^1^ In conclusion, we believe that the performance measures of our models are insufficient to identify persons at high risk of having an UIA, as a first step in the development of screening for UIAs on a population level. Rather, our models provide insight in risk factors contributing to UIA and also show evidence that not only risk factors for aSAH but also for UIAs may have sex-specific effects.^22,28^

UIAs are more prevalent in women than in men, especially after 50 years of age, while SAH incidence in women also increases after that age.^1,2^ It has been hypothesized that lower estrogen levels, estrogen-receptor density, and collagen content of cerebral arteries in women during and after menopause increase the risk of UIA formation and rupture.^29,30^ Also differential effects of hypertension and smoking in men and women on the risk of developing aSAH have been shown.^20,21,27,29,31^ Such effects have not been studied for UIAs yet. Because of these sex differences and the sex-differential effects of risk factors for aSAH, we studied the interactions between sex and all other candidate diagnostic markers. The model of the whole population showed indeed interactions of sex with the diagnostic markers hypertension, and smoking status. This result shows the importance of further research on the differential effect of these risk factors not only for aSAH but also for UIAs. Interestingly, in the model for women smoking status was identified as the only diagnostic marker while the model for men showed more diagnostic markers. Given the relatively low sample size of our study, we cannot draw any definitive conclusions but it could be that yet unknown female-specific risk factors additionally contribute to the risk of UIAs in women.

Hypercholesterolemia and alcohol consumption were no diagnostic markers for prevalent UIAs in the general population. DM was only an independent diagnostic marker of prevalent UIAs in the full study population and not in the models for men and women separately. This does not mean that these factors do not play a role in the risk of UIAs, but rather that they are no diagnostic markers for prevalent UIAs in the general population beyond the other diagnostic markers in the models. Also, other factors may contribute to the risk of having an UIA, such as hormone replacement therapy or family history of stroke or aSAH, but these factors were not available in our study.^19,20^

An important strength of our study is the general population-based study design and sample size that enabled us to study a broad range of candidate diagnostic markers despite the relatively low UIA prevalence. Knowledge on such diagnostic markers in the general population and the construction of a diagnostic model is important as this constitutes the target population for measures to prevent UIA formation. Furthermore, the diagnostic markers in our models are all well-defined and routinely available or easily ascertainable by general practitioners during a standard consultation. We therefore chose not to add, for example, genetic risk factors to our model. Moreover, previous research also has shown that the added value of genetic over clinical data for aSAH and UIAs is limited.^32^

Our study has some limitations. First, the Rotterdam Study assessed presence of UIAs on proton-density T2-weighted brain MRI scans, instead of on MRA with which UIAs are best detected. Moreover, the proton-density T2-weighted brain MRI scans had a slice thickness of 1.6 mm, which means that smaller UIAs may have been missed because of a limited spatial resolution, thereby limiting the power of our study. In a systematic review and meta-analysis from 2011 the overall UIA prevalence was estimated as 3.2% (95%CI 1.9-5.2)^1^ with the UIA prevalence in our study of 2.2% thus being near the lower limit of this 95% CI. Second, participants in the Rotterdam Scan Study were more likely to be white (more than 95% of the study population), middle-class persons. Because of this relatively homogenous study population, generalizability other ethnic or socioeconomic populations may be limited. Third, although we internally validated our models with bootstrapping techniques, our models have not been externally validated, a process recommended to further improve generalizability and applicability.^24,25^ However, as we concluded that the performance measures of our models are insufficient to allow for their use in clinical practice, such external validation has no added value at present. Fourth, although we judge the performance measures of our models as insufficient, we do not know what the performance of such a diagnostic model should exactly be to achieve a cost-effective screening program. This should ideally be assessed in a cost-effectiveness study. Last, preferably we would like to further assess the consistency of our findings in other, larger, population cohorts with data on the presence of UIAs but to our knowledge these are currently not available. We could only identify even smaller populations with 19 persons with UIAs identified in 1006 participants of the Nord-Trøndelag Health (HUNT) Study and 122 persons with UIAs identified in 1862 participants of the Tromsø Study.^33,34^ Future studies are also needed to investigate sex-differential effects of risk factors and potential female specific risk factors for UIAs.

On analyzing diagnostic models for estimating individual persons’ absolute risk of having an UIA in the general population, the diagnostic model separately for men had the best performance. Despite this model showing the best performance, the sensitivity and specificity of the model for UIA presence was limited and the model could not identify men with an UIA prevalence above the average UIA prevalence in the general population. Therefore, this model cannot contribute to identify individuals at high risk of UIAs in the general population who might be eligible for screening. Larger population cohorts with data on the presence of UIAs are needed to assess the consistency of our findings. Future research should also further assess sex-differential effects of risk factors and explore potential female specific risk factors for UIAs. Last, cost effectiveness studies are necessary to determine the exact performance measures of a diagnostic model to achieve a cost-effective screening program for UIAs in the general population.

## Data Availability

The data that support the findings of this study are available from the corresponding author upon reasonable request.

## Acknowledgements

We gratefully acknowledge the study participants of the Ommoord district and their general practitioners and pharmacists for their devotion in contributing to the Rotterdam Study. We also thank all staff who facilitated assessment of participants in the Rotterdam Study throughout the years.

